# Building a prognostic tool for disorders of consciousness: protocol for a multimodal imaging study (IMAGINA study)

**DOI:** 10.1101/2023.01.19.23284810

**Authors:** Beaudoin-Gobert Maude, Merida Inès, Costes Nicolas, Perrin Fabien, André-Obadia Nathalie, Dailler Frédéric, Lartizien Carole, Riche Benjamin, Maucort-Boulch Delphine, Luauté Jacques, Gobert Florent

## Abstract

**Background:** In the last decades, advances in Intensive Care Unit management have led to decreased mortality. However, significant morbidity remains as patients survive after a lesional coma with uncertain quality of awakening and high risk of functional disability. Predicting this level of recovery but also the functional disability of those who will awake constitutes a major challenge for medical, ethical and social perspectives. Among the huge heterogeneity of coma-related injuries, recognising the universality of a common functional pattern which may be focused on a final step of an integrated network would be of great interest for our understanding of disorders of consciousness. The objective of this study is to investigate the neural correlates of arousal and awareness in coma and post-coma to build a prognostic tool based on the detection of a common pattern between patients with a favourable *versus* an unfavourable outcome.

**Method/Design:** We will implement this objective in a translational approach which combines PET-MR imaging, neurophysiology, behavioural/clinical assessments and innovative statistical and computational analysis tools in patients with disorders of consciousness in Intensive Care Unit and in Rehabilitation Department.

## INTRODUCTION

### Context

In the last decades, advances in Intensive Care Unit (ICU) management have led to decreased mortality. However, significant morbidity remains as patients survive after a lesional coma with uncertain quality of awakening and high risk of functional disability. The consciousness impairment is classically described in the double dimension of arousal (or level of consciousness, also named wakefulness) and awareness (or content of consciousness)^1^. A partial consciousness recovery is responsible for an evolution toward several kind of disorders of consciousness (DOC): the reappearance of arousal (attested by eye opening periods) defines the vegetative state (or Unresponsive Wakefulness Syndrome, referred in this document as VS/UWS) whereas any clue of a more complex behaviour clinically defined the minimally conscious state (MCS)^2^ when it remains fluctuating and partial (without stable communication). As the classification is currently challenged by a new definition of the MCS category using the concept of Cortically Mediated State inducing a new classification of DOC^3^, we will refer to this group as MCS/CMS.

Predicting this level of recovery but also the functional disability of those who will awake constitutes a major challenge for medical, ethical and social perspectives. Before identifying biomarkers of awakening abilities, the neural mechanisms of consciousness should be pinpointed. Many imaging studies have been conducted in patients with disorders of consciousness (DOC) to explore the neuronal correlate of consciousness. However, the current correlation approach (between the observed heterogeneity of lesion and clinical status) should be overtaken to look for a model of causal inferences between specific lesional patterns and their behavioural consequences.

The brainstem reticular formation is involved in arousal but its integration with the thalamo-cortical network of awareness is not completely understood^4^. The implication of diencephalic structures in pathological fluctuations of arousal is still discussed in their mechanism and possible modulation^5^. Many imaging studies have highlighted the impairment of key subcortical structures in DOC and have led to the “mesocircuit hypothesis”^6^. This hypothesis involves striato-pallidal structures^7^ and the central part of the thalamus^8^ (including intralaminar nuclei relaying arousal inputs from the brainstem). It has also been extensively investigated in the consciousness process and theorized in the “global neuronal workspace hypothesis”^9^ and more recently in the “integrated information theory”^10^. Imaging studies of early cortical thickness^11^ reduction and impairment of functional interaction^12^ point out areas involved in the default mode network (e.g. medial prefrontal and posterior cingulate cortex) or salience and executive control networks^13^. The analysis of specific interactions of injured brainstem reticular formation during coma has recently highlighted the connectivity between anterior cingulate and left anterior insula cortices as cortical candidates for the arousal/awareness balance^4^. However, the mesocircuit hypothesis is not opposed to the implication of cortical networks regarding the brain organization in thalamo-cortical loops^14^. Among these different levels of integration in the conscious process, the thalamus appears to play a key role. Thus, the slight behavioural improvement of a patient in chronic VS/UWS induced by vagus nerve stimulation was associated (but not certainly causally related) with an enhanced metabolic signal in ^18^F-FDG PET in thalamus^5^. In addition, the existence of coma related to bithalamic infarct^15^ emphasizes the role of central thalamus (including the intralaminar nuclei) as a possible place of global integration as it is an anatomical relay of the ascending reticular system input^16^. Even if the thalamus is the output nuclei of the mesocircuit system^8^, its precise contribution remains uncertain. In fact, the thalamic involvement might be sufficient, but not necessary, to alter consciousness. Among the huge heterogeneity of coma-related injuries, recognising the universality of a common functional pattern (independently of specific thalamic lesions^8^), which may be focused on a final step of an integrated network would be of great interest for our understanding of DOC.

### Study objectives

The objective of this study is to investigate the neural correlates of arousal and awareness in coma and post-coma. We will implement this objective in a translational approach which combines PET-MR imaging, neurophysiology, behavioural/clinical assessments and innovative statistical and computational analysis tools in patients with disorders of consciousness. The aim is to unravel the pivotal role of the thalamus and define its core function in interaction with other deep structures within the mesocircuit and cortical networks. We expect i) identifying key candidates structures associated with arousal and responsiveness, iii) implementing clinical data to develop a prognostic tool using every available data to predict patients’ outcome.

## METHODS

### Study design

We will manage a clinical study on a complete multimodal dataset (PET-MR imaging, standard EEG, EPs, ERPs and combined connectivity metrics) to assess in acute comatose patients from ICU and chronic DOC the correlation between these recordings and patients’ prognosis or diagnosis, respectively.

### Population and recruitment

We will recruit: i) 40 acute comatose patients presenting a complete/multidimensional behavioural loss of consciousness; ii) 20 chronic patients with DOC exhibiting miscellaneous arousal/awareness impairments patterns (VS/UWS and MCS/CMS) and iii) 25 matched (in sex and age) healthy subjects to constitute a normative database (10 for a PET-MRI normative database, 15 for MRI normative database).

Acute comatose patients should present a lesional cause of coma (traumatic, vascular, anoxic) with no response to simple command at least 48h after sedation’s cessation. They will be included in our study between 7 days and 30 days after the coma onset.

Chronic patients with DOC should present an impairment of awareness (no communication abilities or no functional object manipulation signing an exit MCS/CMS classification according to the CRS-R) more than 3 months after an anoxic brain injury or more than 1 year after a non-anoxic one. They will be included during a dedicated hospitalization for clinical evaluation or follow-up.

### Procedure

#### PET-MR imaging

The PET-MR acquisition will be performed at the CERMEP imaging center. The following PET-MR sequences will be used: metabolic PET ([^18^F]FDG), perfusion (ASL), structural MRI (3DT1 for quantitative morphometry and 64-dir DTI) and functional MRI (two resting state sessions at the beginning and at the end of acquisition to assess fluctuations) (Figure 1). In addition, we will also acquire structural MRI necessary for the radiological description of the lesions’ anatomy and for the assessment of lesional load (i.e. the 3DT1, 3DFLAIR, T2*, SWI).

**Figure 1:**
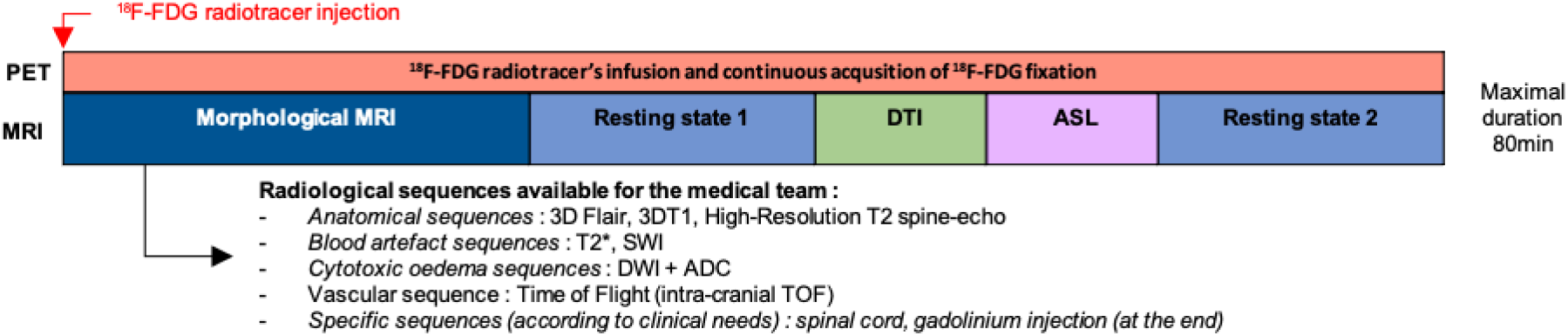
Experimental PET-MR design.

#### EEG

Neurophysiological analysis including EEG can be applied at bedside and operated by the clinical staff. Their prognostic value has been recognized for years after works developed by a local long-standing expertise in ICU and post-intensive rehabilitation unit^27, 29, 30, 74, 75^. Two neurophysiological analyses will be concomitantly performed for comparison within 12h before/after PET-MR acquisition, along with behavioural assessment of the Glasgow Coma Scale (GCS) and the CRS-R:

1. a clinical 20-min EEG (unblinded), along with standard on-line acquisition of EPs (sub-cortical response such as BAEPs and cortical responses such as VEPs, MLAEPs, SEPs) and ERPs (N100, MMN, Subject Own-Name P300).
2. a high-density EEG (128 electrodes) acquired for 30 minutes along with specific ERPs acquisition for off-line analysis (MMN, P3b, Subject Own-Name P300^17^, modulation of SON P300 to music^18^).

#### Pronostic tool

We will use the multi-parametric clinical database to test the synergistic effect of each parameter to reliably predict the prognosis (for acute comatose patients) and refine the accurate diagnosis (for chronic DOC patients). The main task will consist in building a prognostic tool including miscellaneous parameters (clinical, neurophysiological, morphological and functional data using PET and multimodal MRI) for the prediction of awakening at 1 year after a traumatic or vascular brain injury aiming at the optimal reduction of a “functional grey zone” in case of incomplete awakening or reduced autonomy. This functional grey zone will be studied thanks to the outcome at 1-year using a differential dichotomization to build an overall score as the composite of: i) a sub-score predicting a pejorative functional evolution (death, VS/UWS and MCS/CMS); ii) a sub-score predicting the awareness recovery (MCS/CMS, recovery), respectively used as a stringent/liberal criteria allowing several interpretations of awakening.

### Ethics statements

A Consent Form, describing the clinical study and approved by the ethic authorities, will be given to patients’ relatives (in particular the trusted person – in French “personne de confiance” – if previously designated). Families’ patients will be given time to decide if they wish to take part in the study and discuss anything with other family members and intensivists in the ICU. Patients’ relatives will be invited to sign and date the consent form. Family’s patients may feel obliged to participate in this study. To avoid this, they will be assured that neither a refusal nor their participation will interfere with the medical care. This study was approved by French ethical committee (CPP Nord Ouest II; NCT04575454).

## Data Availability

All data produced in the present study are available upon reasonable request to the authors

## Fundings

This study is funded by the Agence Nationale pour la Recherche (grant number ANR-18-CE15-0012).

## Notes

### Competing Interest Statement

The authors have declared no competing interest.

### Author Declarations

French ethics committee gave ethical approval for this work (CPP Nord Ouest II; NCT04575454)

## References

1. Laureys, S. The neural correlate of (un)awareness: lessons from the vegetative state. Trends Cogn. Sci. 9, 556–559 (2005).

2. Giacino, J. T. et al. The minimally conscious state: definition and diagnostic criteria. Neurology 58, 349–353 (2002).

3. Naccache, L. Minimally conscious state or cortically mediated state? Brain J. Neurol. 141, 949–960 (2018).

4. Fischer, D. B. et al. A human brain network derived from coma-causing brainstem lesions. Neurology 87, 2427–2434 (2016).

5. Corazzol, M. et al. Restoring consciousness with vagus nerve stimulation. Curr. Biol. CB 27, R994–R996 (2017).

6. Schiff, N. D. Recovery of consciousness after brain injury: a mesocircuit hypothesis. Trends Neurosci. 33, 1–9 (2010).

7. Lutkenhoff, E. S. et al. Thalamic and extrathalamic mechanisms of consciousness after severe brain injury. Ann. Neurol. 78, 68–76 (2015).

8. Fridman, E. A., Beattie, B. J., Broft, A., Laureys, S. & Schiff, N. D. Regional cerebral metabolic patterns demonstrate the role of anterior forebrain mesocircuit dysfunction in the severely injured brain. Proc. Natl. Acad. Sci. U. S. A. 111, 6473–6478 (2014).

9. Noirhomme, Q. et al. Brain connectivity in pathological and pharmacological coma. Front. Syst. Neurosci. 4, 160 (2010).

10. Tononi, G., Boly, M., Massimini, M. & Koch, C. Integrated information theory: from consciousness to its physical substrate. Nat. Rev. Neurosci. 17, 450–461 (2016).

11. Silva, S. et al. Brain Gray Matter MRI Morphometry for Neuroprognostication After Cardiac Arrest. Crit. Care Med. 45, e763–e771 (2017).

12. Silva, S. et al. Disruption of posteromedial large-scale neural communication predicts recovery from coma. Neurology 85, 2036–2044 (2015).

13. Sair, H. I. et al. Early Functional Connectome Integrity and 1-Year Recovery in Comatose Survivors of Cardiac Arrest. Radiology 162161 (2017) doi:10.1148/radiol.2017162161.

14. Alexander, G. E., DeLong, M. R. & Strick, P. L. Parallel organization of functionally segregated circuits linking basal ganglia and cortex. Annu. Rev. Neurosci. 9, 357–381 (1986).

15. Honig, A. et al. Acute bithalamic infarct manifesting as sleep-like coma: A diagnostic challenge. J. Clin. Neurosci. 34, 81–85 (2016).

16. Yeo, S. S., Chang, P. H. & Jang, S. H. The ascending reticular activating system from pontine reticular formation to the thalamus in the human brain. Front. Hum. Neurosci. 7, 416 (2013).

17. Perrin, F. et al. Brain response to one’s own name in vegetative state, minimally conscious state, and locked-in syndrome. Arch. Neurol. 63, 562–569 (2006).

18. Heine, L. et al. Exploration of Functional Connectivity During Preferred Music Stimulation in Patients with Disorders of Consciousness. Front. Psychol. 6, 1704 (2015).

